# Changes in the rate of cardiometabolic and pulmonary events during the COVID-19 pandemic

**DOI:** 10.1101/2021.02.17.21251812

**Authors:** Alex J Walker, John Tazare, George Hickman, Christopher T Rentsch, Elizabeth J Williamson, Krishnan Bhaskaran, David Evans, Kevin Wing, Rohini Mathur, Angel YS Wong, Anna Schultze, Sebastian CJ Bacon, Chris Bates, Caroline E Morton, Helen J Curtis, Emily Nightingale, Helen I McDonald, Amir Mehrkar, Peter Inglesby, Brian MacKenna, Jonathan Cockburn, William J Hulme, Harriet Forbes, Caroline Minassian, Richard Croker, John Parry, Frank Hester, Sam Harper, Rosalind M Eggo, Stephen JW Evans, Liam Smeeth, Ian J Douglas, Laurie Tomlinson, Ben Goldacre

## Abstract

**Background:** There has been extensive speculation about the relationship between COVID-19 and various cardiometabolic and pulmonary conditions. This a complex question: COVID-19 may cause a cardiometabolic or respiratory event; admission for a clinical event may result in hospital-acquired SARS-CoV-2 infection; both may contribute to a patient surpassing the threshold for presenting to services; and the presence of a pandemic may change whether patients present to services at all. To inform analysis of these questions, we set out to describe the overall rate of various key clinical events over time, and their relationship with COVID-19.

**Methods:** Working on behalf of NHS England, we used data from the OpenSAFELY platform containing data from approximately 40% of the population of England. We selected the whole adult population of 17m patients and within this identified two further mutually exclusive groups: patients who tested positive for SARS-CoV-2 in the community; and patients hospitalised with COVID-19. We report counts of death, DVT, PE, ischaemic stroke, MI, heart failure, AKI and diabetic ketoacidosis in each month between February 2019 and October 2020 within each of: the general population, community SARS-CoV-2 cases, and hospitalised patients with COVID-19. Outcome events were defined using hospitalisations, GP records and cause of death data.

**Results:** For all outcomes except death there was a lower count of events in April 2020 compared to April 2019. For most outcomes the minimum count of events was in April 2020, where the decrease compared to April 2019 in events ranged from 5.9% (PE) to 40.0% (heart failure). Despite hospitalised COVID-19 patients making up just 0.14% of the population in April 2020, these patients accounted for an extremely high proportion of cardiometabolic and respiratory events in that month (range of proportions 10.3% (DVT) to 33.5% (AKI)).

**Interpretation:** We observed a substantial drop in the incidence of cardiometabolic and pulmonary events in the non-COVID-19 general population, but high occurrence of COVID-19 among patients with these events. Shortcomings in routine NHS secondary care data, especially around the timing and order of events, make causal interpretations challenging. We caution that the intermediate findings reported here should be used to inform the design and interpretation of any studies using a general population comparator to evaluate the relationship between COVID-19 and other clinical events.

## Introduction

High prevalence of cardiometabolic and pulmonary complications has been reported in people with COVID-19^1–3^. The relationship between COVID-19 and cardiometabolic or respiratory events is complex: COVID-19 may cause a cardiometabolic or respiratory event; admission for a clinical event may result in hospital-acquired SARS-CoV-2 infection; both may contribute to admission and diagnosis; and the presence of a pandemic may change whether patients present to services at all, as evidenced by a substantial drop in the total number of admission for some conditions, such as acute coronary syndromes^4^.

We set out to describe the overall rate of various key clinical events over time, and their relationship with COVID-19, using the linked data within OpenSAFELY including detailed primary care records, hospital episodes and spells from SUS, and ONS death certificates. Our aim was to provide a broad overview of how the rate of recorded outcome events has changed during the pandemic across the whole population; and to describe the co-occurrence of COVID-19 and each clinical event, in order to inform the design and interpretation of future studies seeking to understand causal relationships with COVID-19.

## Methods

### Study design and data sources

We conducted a time-series study, calculating a series of period prevalences, for each month. We used electronic health record (EHR) data from primary care practices using TPP software linked to Office for National Statistics (ONS) death registrations and Secondary Uses Service (SUS) data (containing hospital records) through OpenSAFELY. This is a data analysis platform developed during the COVID-19 pandemic, on behalf of NHS England, to allow near real-time analysis of pseudonymised primary care records at scale, covering approximately 40% of the population in England, operating within the EHR vendor’s highly secure data centre.^5,6^ Details on Information Governance for the OpenSAFELY platform can be found in the Appendix.

### Population

We extracted monthly cohorts containing the whole adult population (age ≥18 years) who were alive on the first day of each month (February 2019 to October 2020 inclusive, the months for which there was complete data). The population was stratified by presence or absence of a positive SARS-CoV-2 test result, as well as being hospitalised with COVID-19. All people were in the general population group unless they had a positive test for SARS-CoV-2 or were hospitalised with COVID-19. People were categorised in the SARS-CoV-2 positive group if they had a positive test in that month, or in any previous month. Similarly, people were categorised into the hospitalised with COVID-19 group if they were hospitalised with COVID-19 during that month, or in any previous month. If a person both tests positive and is hospitalised with COVID-19, then they are categorised into the hospitalised with COVID-19 group from the month that the COVID-19 hospitalisation occurs.

### Outcomes

We measured eight outcomes: deep vein thrombosis (DVT), pulmonary embolism (PE), ischaemic stroke, myocardial infarction (MI), heart failure, acute kidney injury (AKI) and ketoacidosis. Outcomes were defined as the presence of a diagnostic code for each of the respective outcomes, either in the general practice record, in hospital, or as a cause of death on a death certificate. For AKI and ketoacidosis, the outcomes were restricted just to events recorded in hospital or on the death certificate.

### Statistical methods

For each month, we counted the number of patients with each of the relevant outcome events. Each person was counted only once each month, but people could appear in multiple months if they have repeated records of the outcome. Counts where there were fewer than 5 people in a strata were redacted to ensure anonymity. We calculated 95% confidence intervals for the total counts for each month.

### Software and reproducibility

Data management was performed using the OpenSAFELY software, Python 3.8 and SQL, and analysis using Python 3.8. All codelists alongside code for data management and analyses can be found at: github.com/opensafely/population-outcomes-burden-research. All software for the OpenSAFELY platform is available for review and re-use at github.com/opensafely-core.

## Results

The base population was similar across all months, with an average population of 17,595,257 adults. The SARS-CoV-2 positive population started at zero before the pandemic, before increasing to 209,752 people by October 2020. The population hospitalised with COVID-19 reached 32,372 by October 2020.

Counts of each outcome per month are presented in Figure 1. All outcomes show a decrease in events during the pandemic, with the exception of death, where there was an increase, and ketoacidosis, where uncertainty is higher due to the smaller number of events. Full tables containing the counts on which Figure 1 is based, as well as figures for the base population can be found at github.com/opensafely/population-outcomes-burden-research/tree/master/released_output.

**Figure 1:**
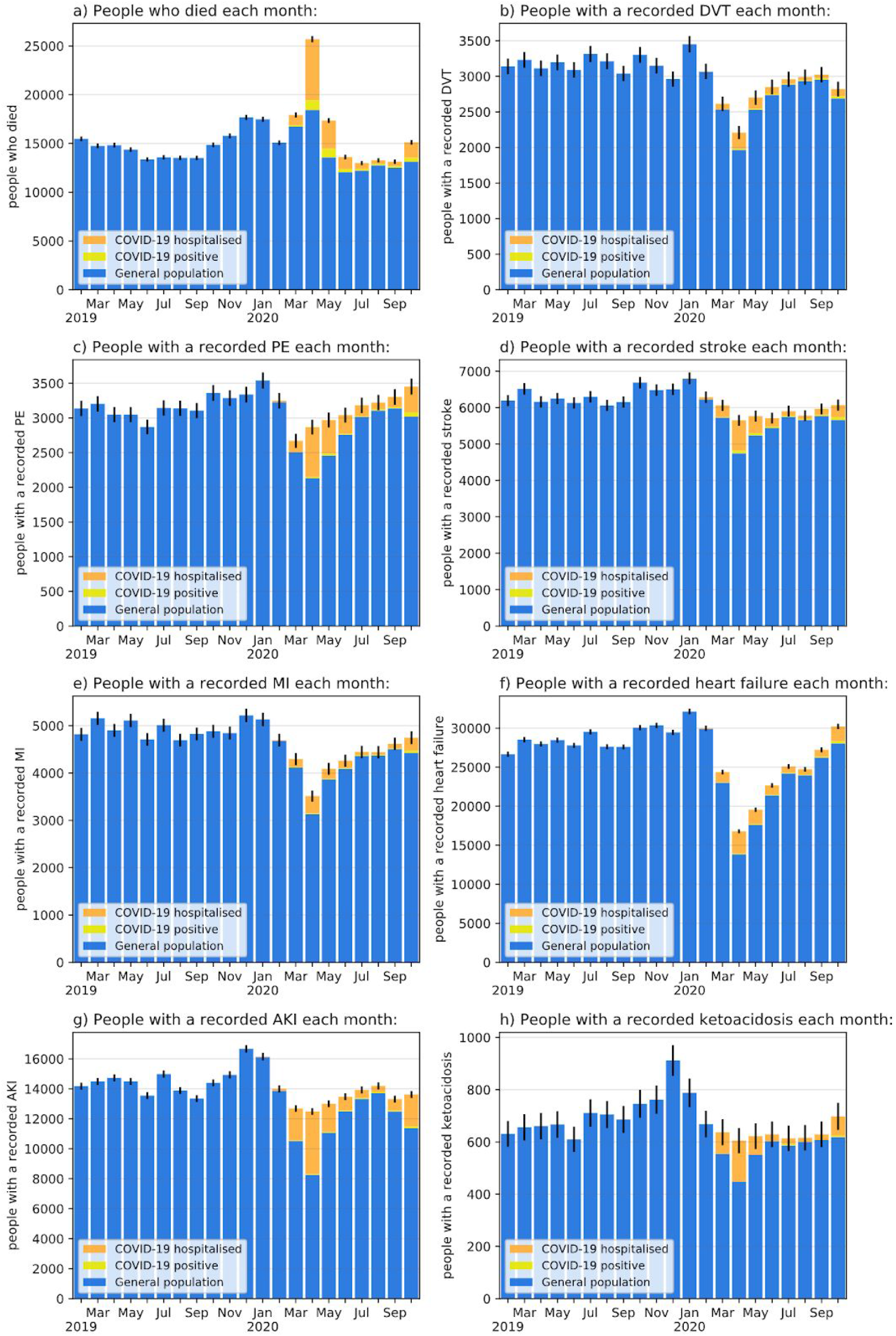
Counts of people with adverse events in the whole population. Coloured strata in each column represent the general population (blue), those testing positive for SARS-CoV-2 in that month or any time before (yellow), and those hospitalised with COVID-19 during that month or any time before (orange). Error bars represent 95% confidence intervals. Note that the number of events for each condition (on the y-axis scale) varies substantially.

During the peak of the first wave of the pandemic, in April 2020, total deaths increased by 73.4% compared to a year earlier (April 2019), while totals for all other outcomes decreased by between 5.9% and 40.0%. Full counts and changes are in Table 1. Counting only events that occured in patients without a record of SARS-CoV-2 infection or hospitalisation with COVID-19, then reductions are greater still.

**Table 1:**
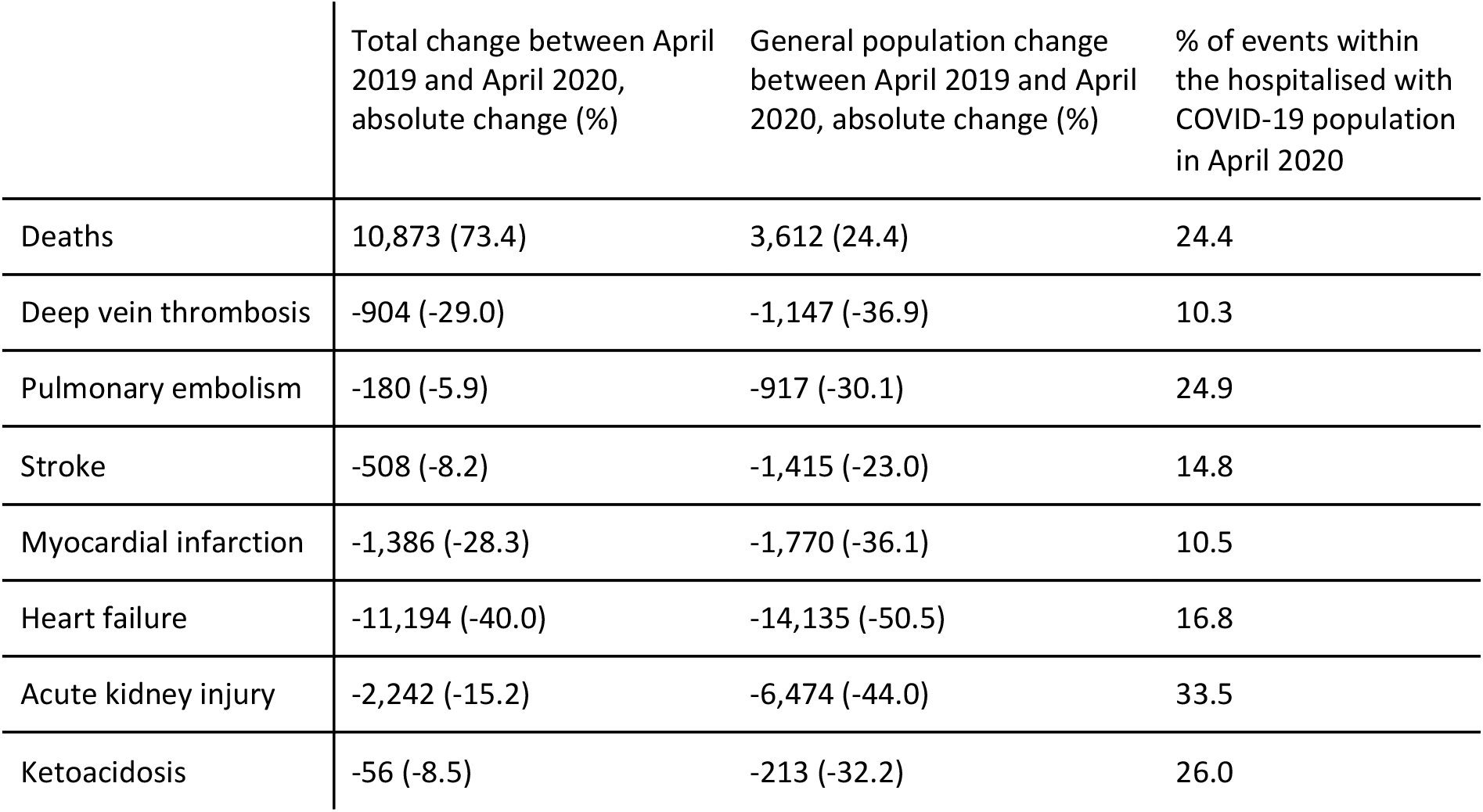
Changes in counts of events between April 2019 and April 2020, as well as the proportion of events occurring in patients recorded as hospitalised with COVID-19 in April 2020.

|  | Total change between April 2019 and April 2020, absolute change (%) | General population change between April 2019 and April 2020, absolute change (%) | % of events within the hospitalised with COVID-19 population in April 2020 |
| --- | --- | --- | --- |
| Deaths | 10,873 (73.4) | 3,612 (24.4) | 24.4 |
| Deep vein thrombosis | -904 (-29.0) | -1,147 (-36.9) | 10.3 |
| Pulmonary embolism | -180 (-5.9) | -917 (-30.1) | 24.9 |
| Stroke | -508 (-8.2) | -1,415 (-23.0) | 14.8 |
| Myocardial infarction | -1,386 (-28.3) | -1,770 (-36.1) | 10.5 |
| Heart failure | -11,194 (-40.0) | -14,135 (-50.5) | 16.8 |
| Acute kidney injury | -2,242 (-15.2) | -6,474 (-44.0) | 33.5 |
| Ketoacidosis | -56 (-8.5) | -213 (-32.2) | 26.0 |

The population of patients hospitalised with COVID-19 was 0.14% of the total population in April 2020, rising to 0.18% by October 2020. Hospitalised COVID-19 patients are dramatically overrepresented in the population of patients who had outcome events, gIven that the proportion of events occuring in this population ranges from 10.3% to 33.5%.

## Discussion

### Summary

Our study has two key findings that are superficially in conflict with each other: a range of key cardiometabolic and respiratory events were substantially more common in patients who also had COVID-19; however the incidence of these conditions in the whole population overall fell substantially during the peak of the COVID-19 pandemic in early 2020, with the reduction in event rates varying between 5.9% and 40% for different outcomes. The proportion of cardiometabolic and respiratory events where the patient was also hospitalised with COVID-19 varied between 10.3 and 33.5%.

### Strengths and limitations

A key strength of our analysis is size: our analysis is based on the detailed primary care and summary hospital records of over 17 million people. Our aim was to give a high level overview of how the volume of events changed during the pandemic, and the co-occurrence of COVID-19 and each event: although we observed a high proportion of events within the COVID-19 population, we did not design the study to demonstrate a causal link between the two. Such relationships are likely to be complex: for example, some patients may be admitted with a cardiometabolic or respiratory event, and then acquire COVID-19 within hospital; some patients may develop COVID-19 and then experience a cardiometabolic or respiratory event as a consequence; some patients may have both COVID-19 and a cardiometabolic or respiratory event in the community, with both contributing to their presentation to services. We did not aim to determine the order that events occurred in: however in many cases the order of events cannot be reliably ascertained from the data available at national level, such as SUS or HES data, combined with SGSS test date data. For example it is not possible to reliably determine the time course or order of events within a hospital spell.

### Findings in context

A previous study using in-hospital records reported a 40% reduction in the incidence of acute coronary syndromes during the peak of Wave 1 of the pandemic in England, consistent with our findings, but did not explore co-occurrence with COVID-19^4^. Our definition of myocardial infarction was similar, but also measured events recorded in the community and on death certificates: we report a 28.3% reduction in events. A recent preprint using detailed COVID-19 Clinical Information Network/International Severe Acute Respiratory and Emerging Infection Consortium (ISARIC) data alongside sparse SUS/HES data estimated that 20-25% of SARS-CoV-2 infections were hospital-acquired during the first wave of the pandemic in England^7^.

### Implications and future research

Our results suggest the SARS-CoV-2 pandemic is affecting incidence and recording of events in a variety of ways. COVID-19 appears to be strongly associated with various cardiometabolic and respiratory events; but the rate of those events fell very substantially during the peak of the first wave of COVID-19. The reduction in these cardiometabolic and respiratory events across the population may be driven by a range of factors: patients may have been reluctant to present to services with cardiometabolic and respiratory events due to fear of infection during the peak of the pandemic; or there may have been a true reduction in incidence of those events, perhaps caused by a change in exposure to risk factors such as air pollution or strenuous activity; or multiple factors may have combined to produce the reduction in presentations. Competing risk of death is also likely to be important, as many of those who died from COVID-19 would also be at high risk of the measured outcomes.

We therefore present this intermediate data for the wider community to inform study designs and further analyses by our group and others. We suggest two immediate future directions: firstly, more complex analyses of existing national data, consisting of detailed primary care data, and sparse secondary care data such as SUS and HES where the timing and order of events is unclear; secondly, further analysis of timing and order of events in richer datasets covering a smaller population, such as that managed by ISARIC^8^, or other bespoke data collections. We caution that the intermediate findings reported here should be used to inform the design and interpretation of any studies using a general population comparator to measure the relationship between COVID-19 and other clinical events. The uncertainties identified in the data, and the results, both highlight the need for greater granularity of data related to details and timing of health events occuring in hospital.

## Data Availability

All data were linked, stored and analysed securely within the OpenSAFELY platform (https://opensafely.org/). Detailed pseudonymized patient data are potentially re-identifiable and therefore not shared. We rapidly delivered the OpenSAFELY data analysis platform without prior funding to deliver timely analyses of urgent research questions in the context of the global COVID-19 health emergency: now that the platform is established, we are developing a formal process for external users to request access in collaboration with NHS England. Details of this process will be published in due course on the OpenSAFELY website.

https://github.com/opensafely/population-outcomes-burden-research

## Acknowledgements

We are very grateful for all the support received from the TPP Technical Operations team throughout this work; for generous assistance from the information governance and database teams at NHS England / NHSX.

## Ethical Approval

This study was approved by the Health Research Authority (REC 20/LO/0651) and by the LSHTM Ethics Board (#21863).

## Appendix

### Information governance and ethics

NHS England is the data controller; TPP is the data processor; and the key researchers on OpenSAFELY are acting on behalf of NHS England. OpenSAFELY is hosted within the TPP environment which is accredited to the ISO 27001 information security standard and is NHS IG Toolkit compliant;^9,10^ patient data are pseudonymised for analysis and linkage using industry standard cryptographic hashing techniques; all pseudonymised datasets transmitted for linkage onto OpenSAFELY are encrypted; access to the platform is via a virtual private network (VPN) connection, restricted to a small group of researchers who hold contracts with NHS England and only access the platform to initiate database queries and statistical models. Pseudonymised structured data include demographics, medications prescribed from primary care, diagnoses, and laboratory measures. No free text data are included. All database activity is logged; only aggregate statistical outputs leave the platform environment following best practice for anonymisation of results such as statistical disclosure control for low cell counts.^11^ The OpenSAFELY research platform adheres to the obligations of the UK General Data Protection Regulation (GDPR) and the Data Protection Act 2018. In March 2020, the Secretary of State for Health and Social Care used powers under the UK Health Service (Control of Patient Information) Regulations 2002 (COPI) to require organisations to process confidential patient information for the purposes of protecting public health, providing healthcare services to the public and monitoring and managing the COVID-19 outbreak and incidents of exposure; this sets aside the requirement for patient consent.^12^ Taken together, these provide the legal bases to link patient datasets on the OpenSAFELY platform. GP practices, from which the primary care data are obtained, are required to share relevant health information to support the public health response to the pandemic, and have been informed of the OpenSAFELY analytics platform. This study was approved by the Health Research Authority (REC reference 20/LO/0651) and by the LSHTM Ethics Board (ref 21863).

## References

1. Chan, L. et al.. AKI in Hospitalized Patients with COVID-19. J. Am. Soc. Nephrol. 32, 151–160 (2021).

2. Ellul, M. A. et al.. Neurological associations of COVID-19. Lancet Neurol. 19, 767–783 (2020).

3. The OpenSAFELY Collaborative et al. Rates of serious clinical outcomes in survivors of hospitalisation with COVID-19: a descriptive cohort study within the OpenSAFELY platform. medRxiv (2021) doi: 10.1101/2021.01.22.21250304.

4. Mafham, M. M. et al.. COVID-19 pandemic and admission rates for and management of acute coronary syndromes in England. Lancet 396, 381–389 (2020).

5. Williamson, E. J. et al.. OpenSAFELY: factors associated with COVID-19 death in 17 million patients. Nature 1–11 (2020).

6. Coronavirus (COVID-19) Research Platform. NHS England https://www.england.nhs.uk/contact-us/privacy-notice/how-we-use-your-information/covid-19-response/coronavirus-covid-19-research-platform/.

7. Scientific Advisory Group for Emergencies. PHE and LSHTM: The contribution of nosocomial infections to the first wave, 28 January 2021. https://www.gov.uk/government/publications/phe-and-lshtm-the-contribution-of-nosocomial-infections-to-the-first-wave-28-january-2021 (2021).

8. Docherty, A. B. et al.. Features of 20 133 UK patients in hospital with covid-19 using the ISARIC WHO Clinical Characterisation Protocol: prospective observational cohort study. BMJ 369, m1985 (2020).

9. NHS Digital. Data Security and Protection Toolkit. 2020. https://digital.nhs.uk/data-and-information/looking-after-information/data-security-and-information-governance/data-security-and-protection-toolkit.

10. NHS Digital. BETA - Data Security Standards. 2020. https://digital.nhs.uk/about-nhs-digital/our-work/nhs-digital-data-and-technology-standards/framework/beta---data-security-standards.

11. NHS Digital. ISB1523: Anonymisation Standard for Publishing Health and Social Care Data. 2020. https://digital.nhs.uk/data-and-information/information-standards/information-standards-and-data-collections-including-extractions/publications-and-notifications/standards-and-collections/isb1523-anonymisation-standard-for-publishing-health-and-social-care-data.

12. Secretary of State for Health-UK Government. Coronavirus (COVID-19): notification to organisations to share information. 2020. https://www.gov.uk/government/publications/coronavirus-covid-19-notification-of-data-controllers-to-share-information.

